# Not One Enclave: Disaggregation and Cardiometabolic Health in Asian Ethnic Enclaves

**DOI:** 10.64898/2026.02.27.26347282

**Authors:** Eun Young Choi, Virginia W. Chang

## Abstract

Many Asian American (AA) subgroups experience disproportionate rates of cardiometabolic (CMB) conditions, yet the contextual drivers of these disparities remain unclear. Little is known about the role of Asian residential segregation, often conceptualized as Asian enclaves, with limited prior work largely ignoring region of origin and nativity. Using six years of population-based survey data from New York City (N>6,000 AAs) linked with multiple sources of community data, we examine how residence in ethnicity-specific enclaves relates to CMB risks (obesity, hypertension, and diabetes), whether these associations differ by nativity, and the extent to which neighborhood socioeconomic conditions, the built environment, social cohesion, and institutional support account for observed associations. Our combined concentration-based and spatial clustering analysis identified five East Asian enclaves and six South Asian enclaves, with no geographic overlap between the two. Logistic regression analyses show that residence in an East Asian enclave was associated with lower odds of obesity (OR=0.63), while residence in a South Asian enclave was linked to higher odds of diabetes (OR=1.42) and hypertension (OR=1.46). These associations were present only among foreign-born individuals. After adjusting for neighborhood characteristics, the lower obesity risk in East Asian enclaves persisted, while elevated risks in South Asian enclaves were partly reduced. Both suggest a role for unmeasured enclave factors, including cultural and food environments. Our findings challenge the view that Asian enclaves are monolithically health-promoting and redirects scholarly attention toward disaggregated approaches to investigating AA health disparities.

## 1. Introduction

There is growing evidence that many subgroups within the Asian American (AA) community experience disproportionate rates of cardiometabolic (CMB) risk^1^. National data from the United States show a notably high prevalence of diabetes and cerebrovascular disease mortality among AAs compared with non-Hispanic White adults, with substantial heterogeneity across subgroups, including higher CMB disease prevalence among South Asian and Southeast Asians than among East Asian adults.^2,3^ However, the drivers of these disparities remain unclear. Research has primarily focused on individual-level factors^4^, leaving a significant knowledge gap regarding the role of context, particularly neighborhood environment. With a growing influx of immigrants since the 1960s, Asian residential segregation, often conceptualized as *Asian ethnic enclaves*, has been increasing in the US^5,6^. In New York City, approximately 30% of AAs reside in ethnic enclaves^7,8^. These percentages are likely to increase as Asian immigrants are projected to grow and be the largest group of all US immigrants by 2055^9^.

While ethnic enclaves can be a critical upstream factor in the CMB health, their health implications remain unclear. Ethnic enclaves are often seen as protective for health, as they can offer social support networks and protection against racism^10,11^. This is in contrast to the fact that racial residential segregation can also function as a fundamental cause of negative health disparities^12^, particularly for Black Americans^13^. It has been argued that the causes underlying Asian segregation differ from that those for black communities^14^, and may reflect personal preferences to a larger degree than structural disadvantages^5^. Even so, the “healthful enclave” perspective cannot be applied uniformly across Asian subgroups, as mechanisms and historical drivers of segregation vary widely based on their migration histories, cultural backgrounds, and racialized experiences in the US^15^. Empirical findings mirror this complexity, reporting highly mixed associations between Asian ethnic enclave residence and health outcomes. Some studies find that Asian enclave residence is associated with worse health, including poorer self-rated mental health^16^ and lower quality of life^17^. Other studies report protective effects, including healthier dietary patterns^18,19^, lower smoking rates^8,20^, and reduced adverse birth outcomes^21^. A third group of studies reports no significant association for self-reported health^5^, physical activity^18^, diabetes and hypertension^7^, and obesity^22^.

Several key limitations contribute to this inconsistency. First, prior work has used a *pan-national* measure for enclaves without differentiating by Asian subgroup^5,23^. This may not accurately capture the influence of enclaves, because many Asian groups have little to no linguistic or cultural similarities. Second, most studies fail to consider nativity^7,17,19,20^. This is a significant limitation given that approximately 70% of AAs are foreign-born^24^. Third, few studies have attempted to understand the multiple mechanisms through which ethnic enclaves relate to CMB outcomes. This makes it difficult to decouple the potential health benefits of co-ethnic social networks from the negative effects of socioeconomic deprivation and poor built environments.

The present study makes several contributions to addressing these gaps. First, it advances the measurement of ethnic enclaves by using ethnic-specific enclave definitions based on ethnicity, moving beyond prior broad pan-Asian categorizations. We focus on East Asian and South Asian enclaves because preliminary analyses indicated that these groups show sufficiently concentrated and spatially distinct residential patterns to support robust enclave characterization, whereas Southeast Asian residential patterns did not meet criteria for defining distinct enclaves. At the individual-level, Southeast Asian individuals were nevertheless included in the analytic sample. Second, it examines how the health implications of enclave residence differ by nativity status, an underexplored but crucial dimension given the large proportion of foreign-born AAs. Third, this study investigates key explanatory pathways by evaluating whether the relationship between enclave residence and CMB outcomes operates through diverse aspects of neighborhood environment, including neighborhood socioeconomic status (SES), social cohesion, built environment, and institutional support for immigrants. The inclusion of the built environment and institutional support is particularly novel contribution, as prior research has largely overlooked these factors. In the following sections, we expand on each of the three areas by providing more detail on rationale and empirical findings from prior research that informs our specific research questions.

## 2. Background

### 2.1. Ethnic-Specific Asian Enclaves and Health

Most existing research relies on pan-ethnic measures of Asian enclaves, which aggregate diverse national-origin Asian groups into a single construct. This oversimplification likely obscure important differences in the socioeconomic, cultural, and institutional characteristics of enclaves. Several studies have noted this as a key limitation^5,7^, stemming from systematic limitations in data collection as well as conventional practices that collapse heterogeneous Asian populations into a singular category^15^. Empirical evidence shows that Asian ethnic subgroups likely form very different neighborhood environments, which in turn shape how ethnic concentration influences health outcomes. Walton^25^ demonstrates that Korean and Chinese neighborhoods in California are more likely to be affluent, resurgent communities associated with better self-reported health, while Filipino and Vietnamese enclaves are disproportionately low-income communities tied to poor health. Similarly, Janevic et al.^26^ document that South Asian enclaves in New York are associated with an elevated risk for gestational diabetes, whereas Chinese enclaves show no such association. These patterns raise our first set of research questions:

> ***Q1a.*** *How do the location and characteristics of ethnic-specific Asian enclaves in New York City differ by ethnicity?*
>
> ***Q1b****. How is residence in an ethnic-specific Asian enclave (vs. non-enclave areas) associated with CMB outcomes (i.e., obesity, hypertension, and diabetes)?*

### 2.2. The Role of Nativity in Shaping Enclave Impacts

Nativity further complicates these dynamics. While enclaves may serve as a robust environment for new arrivals, it may be less salubrious for native-born AAs and established immigrants. Enclaves can reinforce social and economic isolation from mainstream society, and, when coupled with neighborhood-level deprivation, restrict access to healthcare, quality housing, and safe environments^18^. Thus, enclaves may be less beneficial, or even detrimental, for US-born AAs, who may experience constrained upward mobility or increased psychosocial stress within these environments. Morey et al.^27^ show that living in an Asian enclave is protective against discrimination-related stress for recent immigrants but is associated with higher stress for US-born Asians. This challenges the notion that enclaves are monolithically beneficial for health, as socioeconomic disadvantage may outweigh advantages from social capital especially for later-generation residents. These findings motivate our second research question:

> ***Q2****. Does the association between residence in ethnic-specific Asian enclaves and health outcomes differ by nativity status?*

### 2.3. Neighborhood Pathways Linking Enclaves to Cardiometabolic Health

Understanding the pathways through which ethnic enclave residence shapes health is essential for reconciling the mixed findings in previous studies. Prior research has primarily focused on neighborhood SES and social cohesion mechanisms. For example, most inner-city Asian enclaves in New York and Los Angeles show higher poverty, lower educational attainment, and lower occupational status among residents^28,29^. Many studies found that Asian enclaves with higher poverty rates and fewer economic resources are associated with worse health outcomes^21,25,29^. However, not all enclaves share this profile. Other studies highlight the existence of more affluent Asian enclaves where household incomes, educational attainment, and homeownership rates are comparable to those in predominantly white neighborhoods^30,31^. These affluent communities can provide a wide range of resources, such as bilingual healthcare services, ethnic media, and culturally specific educational programs, that enable residents to navigate both mainstream society and co-ethnic networks^32,33^.

Similarly, co-ethnic social capital has been framed as a protective factor that can buffer stress and enhance access to informal health resources^34^. Co-ethnic neighborhoods tend to foster robust social networks, high social cohesion, and shared cultural identities, which can buffer residents from racial discrimination, reduce stress, and facilitate cooperative resource sharing^35^. This enhanced sense of community can encourage healthier behaviors, increase access to culturally appropriate health information and services, and contribute positively to individual health.

However, neighborhood economic and social conditions do not fully capture the broader structural and environmental contexts that shape health in ethnic enclaves. Two additional dimensions, the built environment and institutional support for immigrants, remain underexamined. Studies find that immigrant-dense Chinese neighborhoods suffer from poorer physical environments characterized by reduced access to walkable infrastructure and fewer exercise facilities^17,18^. Moreover, research from California demonstrates that even socioeconomically affluent Asian-majority neighborhoods experience systematic disinvestment in their physical environments. Compared to white-majority neighborhoods with similar income levels, they tend to have significantly less access to park space, green infrastructure, and walkable amenities^36,37^. These patterns highlight that neighborhood SES and the built environment are not always tightly coupled. This disconnect is likely shaped by broader processes of racialized spatial inequality, where minority communities, even those with economic resources, may face infrastructural neglect and underinvestment. In a similar vein, institutional support for immigrants (e.g., local budget allocations for culturally and linguistically appropriate services and immigrant-serving organizations) may play a critical role in shaping health yet remains rarely addressed in studies. These omissions limit our understanding of how ethnic enclaves function as both risks and resources. This leads to our third research question:

> ***Q3****. To what extent are the associations between ethnic-specific Asian enclave residence and health outcomes explained by neighborhood socioeconomic status, the built environment, social cohesion, and institutional support for immigrants?*

## 3. Methods

### 3.1. Data and Sample

Individual-level data are drawn from the New York City Community Health Survey (NYCCHS), an annual, population-based telephone survey of adults residing in all five boroughs of NYC. To maximize the sample size of AAs, we pooled three 2-year datasets spanning 2015 to 2020, the years for which geographic identifiers at the Public Use Microdata Area (PUMA) level are available, via a data use agreement with the NYC Department of Health. The final analytic sample includes 6,825 AAs. Because missingness varies by outcome, sample sizes differ slightly across models: 6,354 for obesity, 6,436 for hypertension, and 6,434 for diabetes. In stratified analyses, we disaggregate the sample by nativity (US-born: n=998; foreign-born: n=5,806) and by ethnicity (East Asian: n=4,568; South Asian: n=1,631; Southeast Asian: n=410).

Community-level data are drawn from multiple sources. To construct measures of ethnic enclaves, we use population counts by racial/ethnic group from the 2015–2019 American Community Survey (ACS) 5-year estimates. Additional neighborhood characteristics were downloaded from NYC Planning’s Community District Profiles, which provide neighborhood socioeconomic indicators and built environment data. Litter basket coverage was retrieved from the NYC Department of Sanitation, via the Environment & Health Data Portal. Finally, data on public funding for immigrant services were obtained from Data2Go.NYC, reflecting allocations by the Department of Youth and Community Development during the 2019–2020 fiscal year.

### 3.2. Measures

#### 3.2.1. Cardiometabolic (CMB) Risk Outcomes

CMB risk outcomes include obesity, hypertension, and diabetes. Obesity is defined based on self-reported height and weight, with a calculated body mass index of 30 kg/m^2^ or higher. Hypertension and diabetes are assessed through self-reports of having ever been diagnosed with these conditions by a health professional.

#### 3.2.2. Ethnic Enclaves

We defined ethnic-specific Asian enclaves at the PUMA level, which was selected as the geographic unit of analysis because they represent the most granular level available in our individual-level dataset. PUMAs are US Census-defined statistical areas constructed from contiguous census tracts with a minimum population of 100,000. In New York City, PUMAs closely approximate Community Districts (CDs), administratively recognized neighborhood units established in the City Charter and widely used for local planning and service delivery. Because PUMAs generally align with CDs, they capture socially and institutionally meaningful neighborhood contexts, making them a reasonable proxy for enclave boundaries in NYC.

Our classification of Asian ethnic enclaves followed a two-step approach. First, we applied a concentration-based definition to identify PUMAs with high proportions of East Asian and South Asian residents. For each PUMA, we calculated the proportion of residents identifying as East Asian and South Asian within the “Asian alone” race category regardless of Hispanic origin. East Asians include Chinese, Japanese, Korean, Okinawan, and Taiwanese individuals. South Asians include Asian Indian, Bangladeshi, Bhutanese, Nepalese, Pakistani, and Sri Lankan individuals. Because the population shares of these groups differ substantially, we adopted a group-specific threshold using the “double share criterion”^38,39^. Under this approach, a PUMA was classified as an ethnic enclave if the proportion of a specific ethnic group was at least *twice* its share in the overall New York City population. This yielded threshold values of 16.4 percent for East Asians (8.2% citywide) and 8.6 percent for South Asians (4.3%).

Second, we refined this classification by further incorporating a second criterion: spatial clustering of underlying census tracts, which are a sub-unit within PUMAs. This approach reflects the idea that ethnic enclaves involve not just demographic concentration, but also geographic coherence, consistent with established definitions of ethnic neighborhoods^28,40^. Specifically, we applied a measure of local spatial clustering, local Moran’s I statistic^40^. Spatial relationships were defined using a first-order queen contiguity matrix, in which census tracts were considered neighbors if they shared a boundary or a vertex. Local Moran’s I was computed using 999 conditional permutation simulations to estimate statistical significance (*p* <.05). A clustered tract was defined as a tract with an above-average proportion of each ethnic group, surrounded by neighboring tracts that also had above-average proportions, suggesting a spatially cohesive pattern of the group. We then returned to the PUMAs identified as enclaves by the double share criterion above and further required that at least 40% (or a minimum of 15) of its census tracts are classified as spatially clustered. In sum, a PUMA was classified as an Asian ethnic-specific enclave only if it satisfied both the double-share concentration threshold and the presence of statistically significant spatial clustering among its underlying census tracts.

#### 3.2.3. Neighborhood Environment

To assess neighborhood SES and built environment features at the PUMA-level, we examined a broad set of indicators and refined them using factor analysis (details in **S Appendix 1**). The analysis supported a two-factor solution: one factor representing socioeconomic disadvantage (poverty rate, rent burden, no bachelor’s degree, unemployment), and another representing the built environment (park access, number of hospitals, public facilities, and litter basket coverage). Factor scores for each construct were generated and used as continuous predictors in regression models. To assess social cohesion, we aggregated five NYCCHS survey items (perceived close-knit neighborhood, neighbors’ willingness to help, trust among residents, shared values, and whether people in the neighborhood get along) at the PUMA level, capturing perceptions of neighborhood connectedness. Factor analysis supported a single-factor solution, and the resulting factor scores were used to represent area-level cohesion. We also included PUMA-level immigrant-targeted public funding, measured in U.S. dollars and defined as the annual amount of public funding allocated to programs supporting immigrant families, including civic education classes, English for Speakers of Other Languages, and immigration application assistance.

#### 3.2.4. Covariates

We control for individual-level sociodemographic characteristics that may confound the association between enclave residence and health status. These include age group (18-24; 25-64; 65+), sex (female vs. male), nativity (native-born vs. foreign-born), Asian ethnic subgroup membership (East Asian; South Asian; Southeast Asian; Other), marital/partner status (married or cohabiting with a partner vs. divorced, widowed, separated, or never married), education (less than high school vs. high school or above), household income (below 200% of federal poverty guideline vs. higher), and health insurance status (insured vs. uninsured).

### 3.3. Analyses

We first generated age-standardized prevalence estimates of health outcomes by Asian ethnic subgroup using NYCCHS data from 2015 to 2020. All estimates were weighted using CHS-provided survey weights and standardized to the age distribution of New York City’s adult population. To identify ethnic-specific Asian enclaves, we applied a two-step process of concentration and spatial clustering analyses for each ethnic Asian subgroup, as described in earlier sections. We then compared characteristics of adults residing inside and outside each type of Asian enclave to assess demographic and socioeconomic differences.

Next, we use logistic regression models to assess the association of ethnic enclaves with each health outcome, controlling for all covariates. We adjust the standard errors for within-cluster (PUMA) correlation to mitigate the risk of underestimating standard errors in single-level models. Each health outcome (obesity, diabetes, and hypertension) is estimated in a separate regression model, with the ethnic enclave variable included as a categorical predictor (non-enclave as the reference category, compared with East Asian and South Asian enclave residence). We first estimate models for the full sample of Asians and then stratify analyses by Asian ethnic subgroup and nativity status to assess potential heterogeneity in associations. To explore potential explanatory pathways, we compare these neighborhood characteristics between non-enclave areas and each enclave type and then assess how incorporating these factors into regression models accounts for the observed enclave-health associations. This strategy provides a clearer understanding of the ways ethnic enclaves may function as both sources of resilience and sources of risk for CMB health among AA populations.

All analyses were conducted using Stata 18.0. We incorporated sampling weights to account for the NYCCHS’s complex survey design, including stratification and clustering. Each 2-year NYCCHS PUMA dataset provides survey weights calibrated to make each 2-year dataset representative of the average NYC adult population during that 2-year period. Because this study focuses on AA subgroups, we combined three consecutive 2-year datasets to increase the sample size and improve the statistical power for subgroup analyses. However, NYCCHS does not provide weights for 6-year pooled data. To address this, we adjusted the survey weights by dividing each dataset’s 2-year weights by 3 to ensures that the pooled sample reflects the average NYC adult population across the six-year period, avoiding inflation of population estimates. This method follows recommended practices for pooling repeated cross-sectional survey data when the goal is to estimate average effects over time rather than annual trends^41^.

## 4. Results

### 4.1. Health Status Among Asian Ethnic Subgroups

**Figure 1** shows clear disparities in health status across AA ethnic subgroups in New York City. East Asians reported the lowest obesity prevalence (6%), which is substantially lower than that of non-Hispanic Whites (19%), while their rates of diabetes (9%) and hypertension (20%) were comparable to those of Whites. South Asians exhibited markedly higher rates of diabetes (22% vs. 7%) and hypertension (30% vs. 22%), with similar obesity rates (16%) relative to Whites. Southeast Asians also showed elevated hypertension prevalence (26%), but relatively lower obesity (11%) and diabetes rates (8%). These patterns highlight significant AA disparities in CMB disease burden.

**Figure 1.**
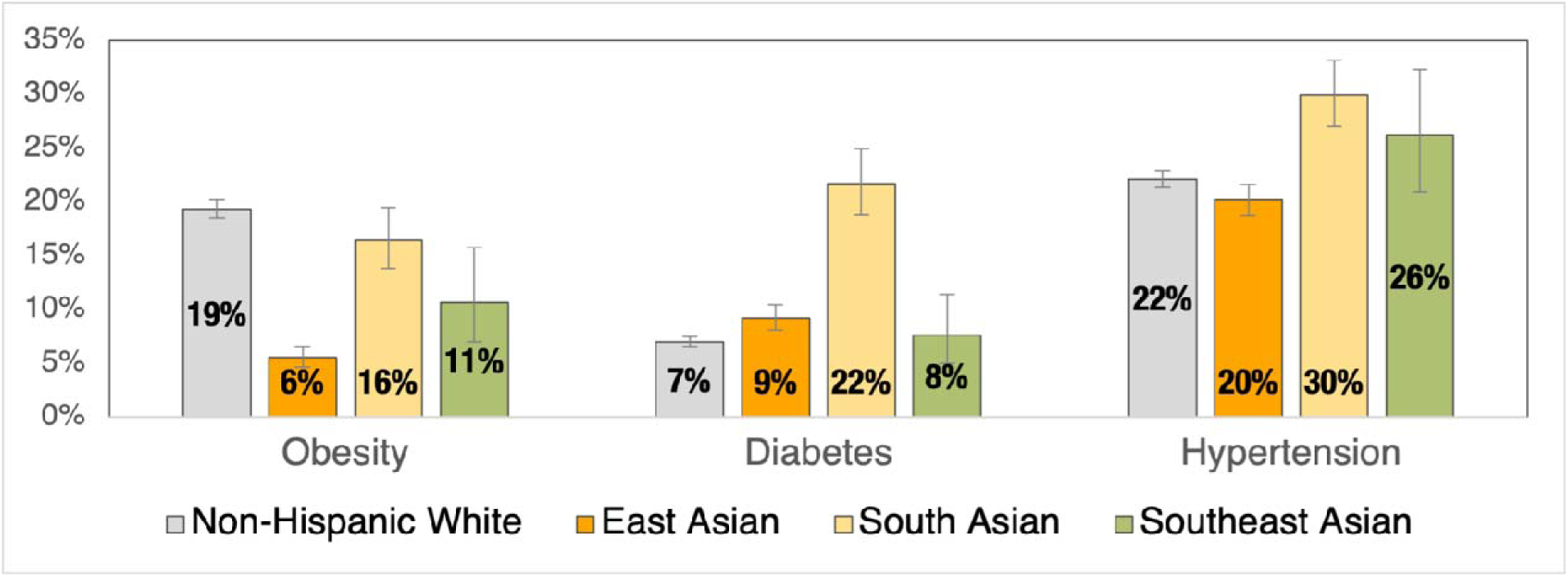
Health Status among Asians in New York City, 2015-2020 NYCCHS *Note.* Estimates were weighted using sample weights and age-standardized using standard age
adjustment weights derived from the age distribution in New York City.

### 4.2. Geographic Distribution of Ethnic-Specific Asian Enclaves

Using the double share concentration threshold combined with the spatial clustering criterion, we identified five East Asian enclaves and six South Asian enclaves. **Figure 2** presents the geographical locations of each Asian enclave in New York City. It shows that there is no geographic overlap between East Asian and South Asian enclaves. East Asian enclaves are primarily located in northeastern Queens and southwestern Brooklyn, including Flushing and parts of Sunset Park. South Asian enclaves are concentrated in southeastern and south-central Queens, in neighborhoods such as Jamaica and Richmond Hill.

**Figure 2.**
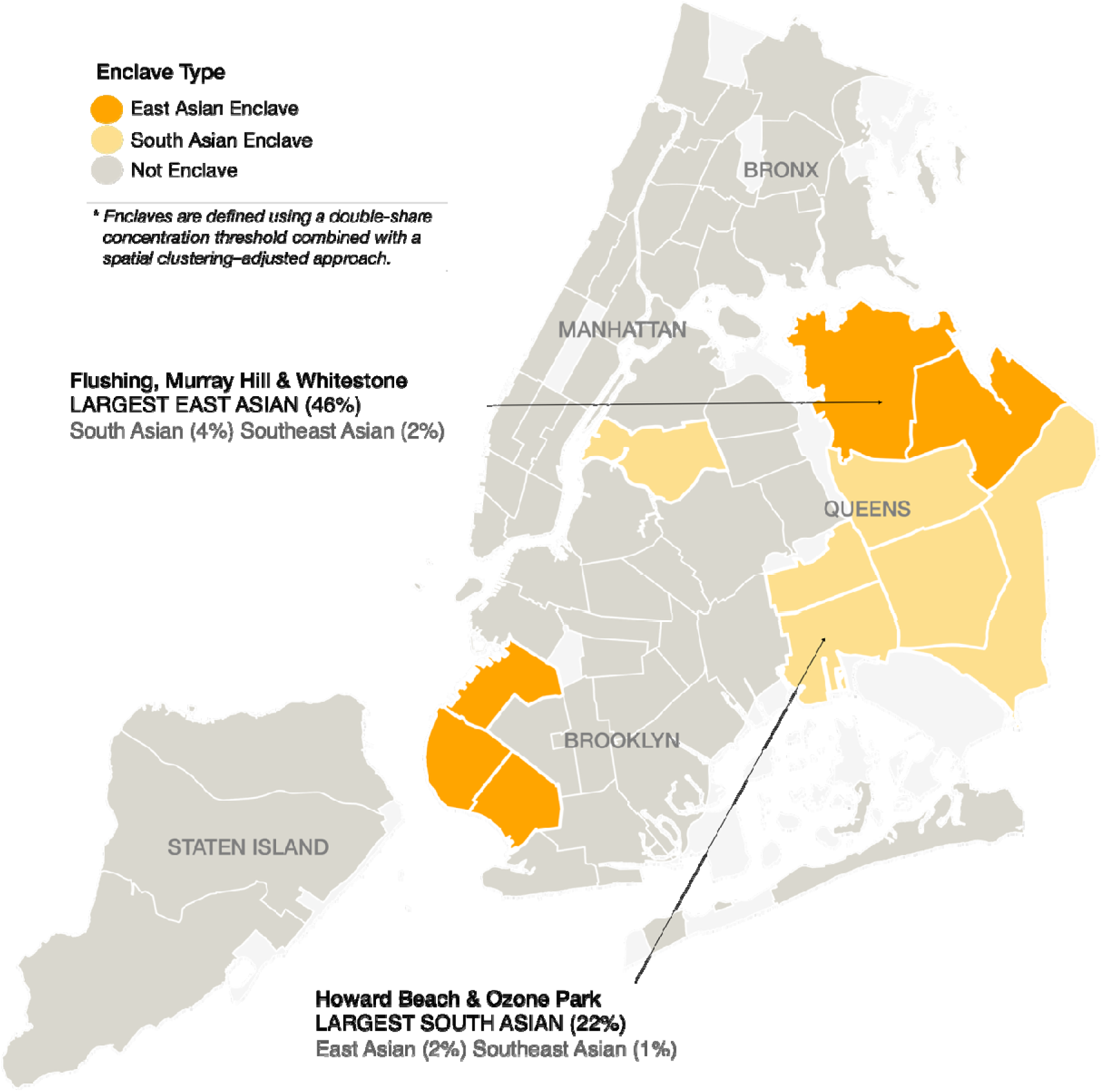
Geographic Locations of Ethnic-Specific Asian Enclaves in New York City *Note.* Light grey areas are joint interest areas such as parks and airports

### 4.3. Racial/Ethnic Composition by Enclave Classifications

**Table 1** presents the racial and ethnic composition of PUMAs across NYC, stratified by enclave classification. As expected, the ethnic composition of enclave-designated PUMAs shows substantially higher proportion of their respective subgroup populations. East Asian enclaves had the highest proportion of East Asian residents (33.9%), compared to 5.1% in non-enclave PUMAs. Similarly, South Asian enclaves had 14.9% South Asian residents, compared to 2.8% in non-enclaves. East Asian and South Asian enclaves also differ in their broader racial and ethnic makeup. East Asian enclaves include a higher proportion of non-Hispanic White residents (36.0% vs. 18.3%), whereas South Asian enclaves have higher shares of non-Hispanic Black (25.0% vs. 2.2%) and Hispanic (24.7% vs. 20.4%) residents. A slightly larger share of Southeast Asian residents lives in South Asian enclaves than in East Asian enclaves (2.8% vs. 1.6%).

**Table 1.** Racial/Ethnic Composition of NYC PUMAs by Enclave Classification.

|  | Non-Enclave | East Asian<br>Enclave | South Asian<br>Enclave |
| --- | --- | --- | --- |
|  | N=44 | N=5 | N=6 |
| Non-Hispanic White | 33.5% | 36.0% | 18.3% |
| Non-Hispanic Black | 23.9% | 2.2% | 25.0% |
| Hispanic | 30.9% | 20.4% | 24.7% |
| Asian category |  |  |  |
| East Asian | 5.1% | 33.9% | 6.9% |
| South Asian | 2.8% | 2.9% | 14.9% |
| Southeast Asian | 1.0% | 1.6% | 2.8% |

### 4.4. Sample Characteristics by Enclave Residency

After linking enclave data to our AA sample, we compared the sociodemographic and health characteristics across enclave residence types. Differences between the reference group and each East Asian and South Asian enclave resident group were assessed with Pearson’s chi-squared tests. As shown in **Table 2**, compared to non-enclave residents, individuals living in East Asian enclaves had significantly lower obesity prevalence (4.2%), while those in South Asian enclaves had notably higher rates of obesity (14.4%), diabetes (15.5%), and hypertension (24.9%). Compared to non-enclave residents, individuals in East Asian enclaves were more likely to be foreign-born. Both East Asian and South Asian enclave residents were more likely to be married or living with a partner. East Asian enclave residents had higher rates of limited education (30.3%) and poverty (69.2%) than non-enclave residents, whereas South Asian enclave residents had lower rates of limited education (16.4%).

**Table 2.** Sample Characteristics by Ethnic-Specific Asian Enclave Residency, All Asians.

|  | Non-enclave<br>Resident<br>(Ref) | East Asian<br>Enclave<br>Resident | South Asian<br>Enclave<br>Resident |
| --- | --- | --- | --- |
| N | 3,651 | 2,030 | 1,144 |
| Obesity | 8.9% | 4.2%*** | 14.4%*** |
| Diabetes | 8.5% | 8.5% | 15.5%*** |
| Hypertension | 17.4% | 18.7% | 24.9%*** |
| Female | 52.7% | 50.5% | 49.3% |
| Foreign born | 83.4% | 90.1%*** | 83.8% |
| Age groups |  |  |  |
| Ages 18-24 | 16.3% | 11.4%*** | 16.1% |
| Ages 25-64 | 72.4% | 76.3%* | 72.1% |
| Ages 65+ | 11.3% | 12.3% | 11.8% |
| Married/living with a partner | 55.5% | 66.5%*** | 61.7%* |
| Less than high school education | 21.5% | 30.3%*** | 16.4%* |
| Households in poverty | 51.8% | 69.2%*** | 53.4% |
| No health insurance | 10.9% | 10.6% | 10.2% |

*Note.* Difference between the reference group and each East Asian and South Asian enclave resident group was assessed with Pearson's chi-squared test. \*\*\* $p < .001$ ; \*\* $p < .01$ ; \* $p < .05$ .

### 4.5. Ethnic-Specific Asian Enclaves and Health Outcomes

**Figure 3** presents the coefficients with 95% CI from logistic regression models estimating the associations between residence in ethnic-specific Asian enclaves and three CMB health outcomes. Separate logistic regression models predicting each obesity, diabetes, or hypertension were estimated by Asian enclave residence status (non-enclaves [reference], East Asian enclaves, and South Asian enclaves). All models adjusted for sociodemographic covariates, including Asian ethnic subgroup membership, which accounts for underlying ethnic differences in individual health status. Living in East Asian enclaves was significantly associated with lower odds of obesity (OR=0.63, *p* <.05), but not with hypertension or diabetes. In contrast, residence in South Asian enclaves was significantly associated with higher odds of hypertension (OR=1.42, *p* <.05) and diabetes (OR=1.46, *p* <.05).

**Figure 3.**
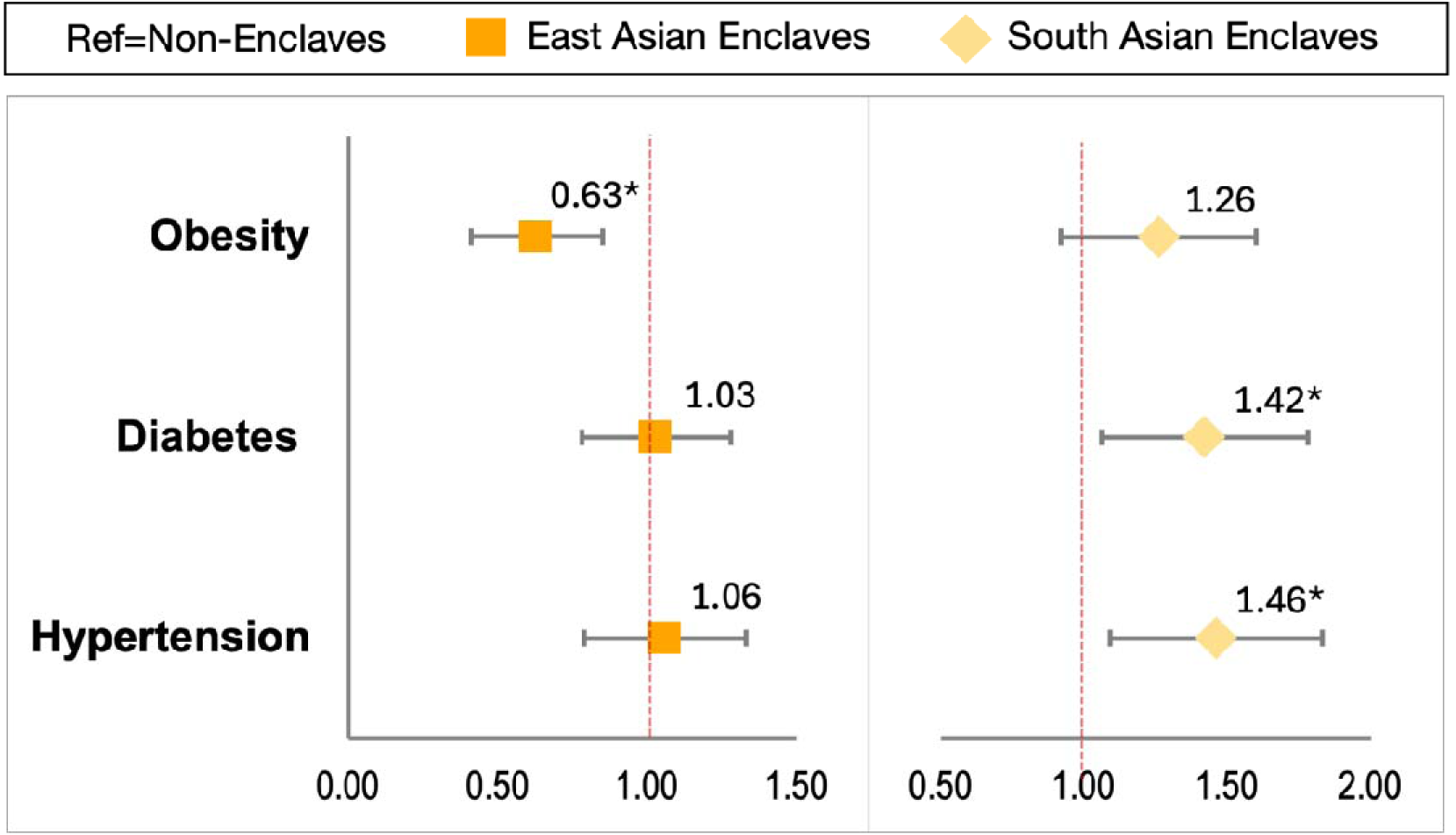
Associations between Asian Enclave Residency and Cardiometabolic Health Outcome *Note.* Estimates represent adjusted odds ratios (ORs) and 95% confidence intervals from separate logistic regression models predicting each obesity, diabetes, or hypertension by Asian enclave residence status (non-enclaves [reference], East Asian enclaves, and South Asian enclaves). Models are adjusted for age, sex, nativity, Asian ethnic subgroup, marital status, education, poverty, and health insurance. * p <.05

Subgroup analyses by Asian ethnic subgroup (**Table 3**) generally mirrored the patterns observed in the full Asian sample. Enclave-health associations were directionally consistent with the pooled results: East Asian enclave residence tended to be associated with lower odds of obesity, while South Asian enclave residence was associated with higher odds of adverse health outcomes. Although some associations did not reach statistical significance within subgroups, likely due to reduced sample sizes, the general protective pattern of East Asian enclaves and the adverse associations of South Asian enclaves remained evident, *regardless of the resident’s own ethnic subgroup*.

**Table 3.** Associations between Ethnic-Specific Asian Enclave and Cardiometabolic Health Outcome by Asian Ethnic Subgroups.

|  | N (%) | Obesity |  |  | Diabetes |  |  | Hypertension |  |  |
| --- | --- | --- | --- | --- | --- | --- | --- | --- | --- | --- |
|  |  | OR |  | 95% CI | OR |  | 95% CI | OR |  | 95% CI |
| *Non-Enclaves (ref)* | 2331 (51%) |  |  |  |  |  |  |  |  |  |
| *East Asian Enclaves* | 1864 (41%) | 0.56 | * | 0.36 0.87 | 1.07 |  | 0.82 1.39 | 1.07 |  | 0.82 1.40 |
| *South Asian Enclaves* | 373 (8%) | 1.00 |  | 0.66 1.49 | 1.60 | *** | 1.24 2.08 | 1.40 |  | 0.89 2.19 |

**B. South Asian**
|  | N (%) | Obesity |  |  | Diabetes |  |  | Hypertension |  |  |
| --- | --- | --- | --- | --- | --- | --- | --- | --- | --- | --- |
|  |  | OR |  | 95% CI | OR |  | 95% CI | OR |  | 95% CI |
| *Non-Enclaves (ref)* | 868 (53%) |  |  |  |  |  |  |  |  |  |
| *East Asian Enclaves* | 90 (6%) | 0.82 |  | 0.43 1.57 | 0.90 |  | 0.33 2.45 | 1.45 |  | 0.51 4.11 |
| *South Asian Enclaves* | 673 (41%) | 1.70 | * | 1.09 2.67 | 1.32 |  | 0.91 1.93 | 1.66 | ** | 1.18 2.33 |

**C. Southeast Asian**
|  | N (%) | Obesity |  |  | Diabetes |  |  | Hypertension |  |  |
| --- | --- | --- | --- | --- | --- | --- | --- | --- | --- | --- |
|  |  | OR |  | 95% CI | OR |  | 95% CI | OR |  | 95% CI |
| *Non-Enclaves (ref)* | 293 (72%) |  |  |  |  |  |  |  |  |  |
| *East Asian Enclaves* | 48 (12%) | 0.67 |  | 0.28 1.60 | 1.13 |  | 0.27 4.78 | 0.92 |  | 0.28 3.02 |
| *South Asian Enclaves* | 69 (17%) | 0.64 |  | 0.21 1.93 | 3.20 | * | 1.07 9.60 | 1.52 |  | 0.68 3.37 |

*Note.* Model was adjusted for age group, sex, nativity, marital/partner status, education, household income, and health insurance status.
\*\*\* $p < .001$ ; \*\* $p < .01$ ; \* $p < .05$

### 4.6. Asian Enclaves and Health Outcomes by Nativity

In nativity-stratified models (**Table 4**), enclave associations were pronounced only among foreign-born Asians. Among them, living in East Asian enclaves was significantly associated with lower odds of obesity (OR=0.54, *p* <.01), and living in South Asian enclaves was associated with higher odds of obesity (OR=1.30, *p* <.01), diabetes (OR=1.41, *p* <.05), and hypertension (OR=1.50, *p* <.01). In contrast, none of the enclave associations reached statistical significance among US-born Asians.

**Table 4.**
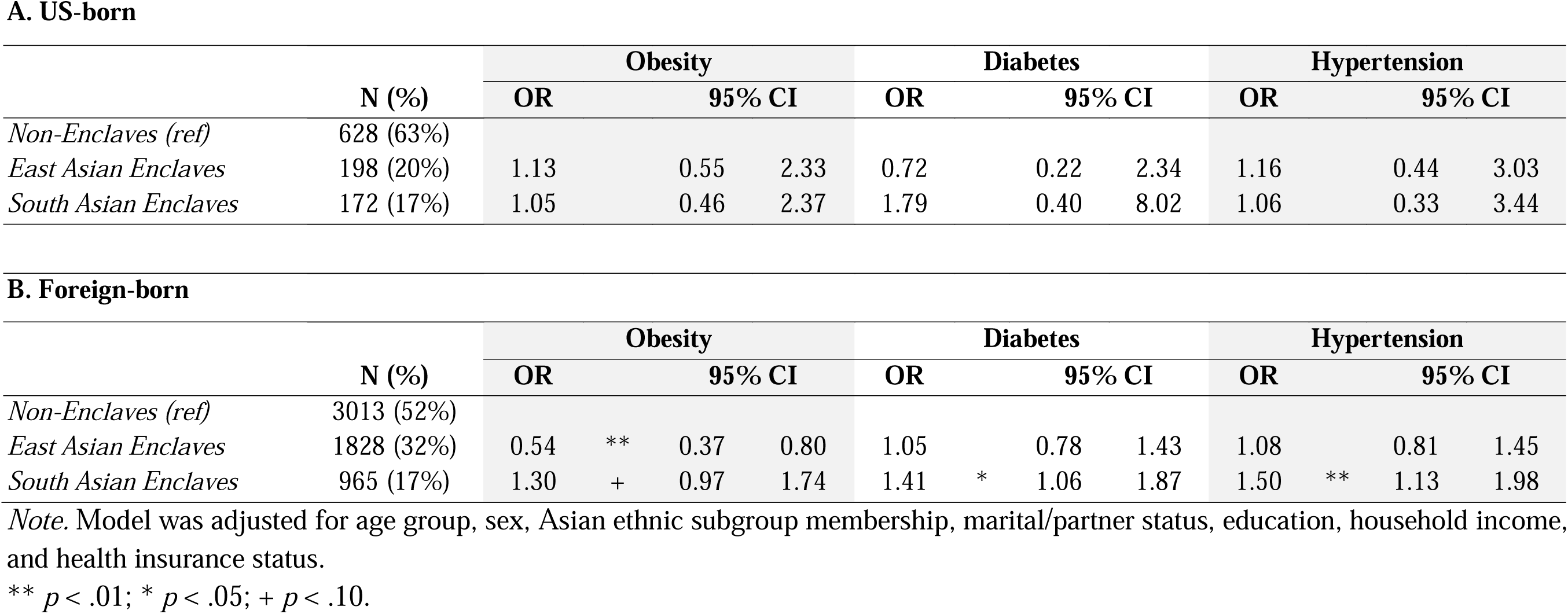
Associations between Ethnic-Specific Asian Enclave and Cardiometabolic Health Outcome by Nativity.

|  | N (%) | Obesity |  |  | Diabetes |  |  | Hypertension |  |  |
| --- | --- | --- | --- | --- | --- | --- | --- | --- | --- | --- |
|  |  | OR | 95% CI |  | OR | 95% CI |  | OR | 95% CI |  |
| *Non-Enclaves (ref)* | 628 (63%) |  |  |  |  |  |  |  |  |  |
| *East Asian Enclaves* | 198 (20%) | 1.13 | 0.55 | 2.33 | 0.72 | 0.22 | 2.34 | 1.16 | 0.44 | 3.03 |
| *South Asian Enclaves* | 172 (17%) | 1.05 | 0.46 | 2.37 | 1.79 | 0.40 | 8.02 | 1.06 | 0.33 | 3.44 |

**B. Foreign-born**
|  | N (%) | Obesity |  |  |  | Diabetes |  |  |  | Hypertension |  |  |  |
| --- | --- | --- | --- | --- | --- | --- | --- | --- | --- | --- | --- | --- | --- |
|  |  | OR |  | 95% CI |  | OR |  | 95% CI |  | OR |  | 95% CI |  |
| *Non-Enclaves (ref)* | 3013 (52%) |  |  |  |  |  |  |  |  |  |  |  |  |
| *East Asian Enclaves* | 1828 (32%) | 0.54 | ** | 0.37 | 0.80 | 1.05 |  | 0.78 | 1.43 | 1.08 |  | 0.81 | 1.45 |
| *South Asian Enclaves* | 965 (17%) | 1.30 | + | 0.97 | 1.74 | 1.41 | * | 1.06 | 1.87 | 1.50 | ** | 1.13 | 1.98 |

*Note.* Model was adjusted for age group, sex, Asian ethnic subgroup membership, marital/partner status, education, household income, and health insurance status.
\*\* $p < .01$ ; \* $p < .05$ ; + $p < .10$ .

### 4.7. Variation in Neighborhood Conditions by Enclave Status

Descriptive comparisons of PUMA-level characteristics (**Table 5**) revealed some differences in neighborhood profiles by Asian enclave type. Compared to non-enclave areas, East and South Asian enclaves had slightly higher levels of socioeconomic disadvantage, with higher rates of rent burden (46-48% vs. 44% in non-enclaves) and a greater proportion of residents lacking a bachelor’s degree (66-70% vs. 63% in non-enclaves). Social cohesion indicators were relatively similar across enclave types and non-enclaves, with only modest differences. Both Asian enclaves had fewer built environment amenities, shown as substantially limited access to a park (57-67% vs. 77% in non-enclaves), fewer number of urgent care facilities (13-17 vs. 24 in non-enclaves), less public institutions (6-7% vs. 11% in non-enclaves), lower litter basket density per square mile (47-61 vs. 171 in non-enclaves), than non-enclave areas. However, notable differences emerged in immigrant-targeted funding: East Asian enclaves received substantially more funding for immigrant services ($418,021), while South Asian enclaves received considerably less ($70,430) than non-enclaves ($245,373).

**Table 5.** Differences in PUMA-level Profiles by Asian Enclave Status.

|  | Non-Enclave | East Asian<br>Enclave | South Asian<br>Enclave |
| --- | --- | --- | --- |
|  | N=44 | N=5 | N=6 |
| **Socioeconomic Status** |  |  |  |
| Residents with incomes below NYC poverty threshold, unit: % | 20.0 | 19.9 | 16.4 |
| HHs spend >35% on rent, unit: % | 44.4 | 48.1 | 46.4 |
| Residents without a bachelor's degree+, unit: % | 62.6 | 65.5 | 70.4 |
| Civilian labor force unemployed, unit: % | 4.6 | 3.4 | 4.7 |
| **Social Environment** |  |  |  |
| Live in a close-knit neighborhood, unit: % agree | 66.1 | 67.6 | 69 |
| Neighbors are willing to help, unit: % agree | 77.0 | 76.0 | 78.4 |
| Neighbors get along with each other, unit: % agree | 70.4 | 69.3 | 74.0 |
| Neighbors share the same values, unit: % agree | 54.3 | 55.9 | 58.1 |
| Neighbors can be trusted, unit: % agree | 62.6 | 68.6 | 67.6 |
| **Built Environment** |  |  |  |
| Residents within walking distance of a park or open space, unit: % | 76.8 | 67.0 | 56.8 |
| Urgent care hospitals, centers, and health facilities, unit: count | 24.2 | 16.6 | 13.2 |
| Area public facilities/institutions as the primary land use, unit: % | 10.7 | 6.9 | 6.1 |
| Litter baskets, unit: density per square mile | 171.4 | 61.3 | 47.4 |
| **Institutional Support for Immigrants** |  |  |  |
| Funding for literacy and immigrant services, unit: \$ | \$ 245,373 | \$ 418,021 | \$ 70,430 |

*Note.* Public Facilities and Institutions include hospitals, churches, Asylums and homes, educational structures, community center, etc.

### 4.8. Community-Level Pathways Linking Asian Ethnic-Specific Enclaves to Health

Given that enclave–health associations were observed only among foreign-born AAs, we estimated models for this subgroup that simultaneously incorporated all neighborhood socioeconomic, social, built, and institutional factors to examine whether the enclave associations could be explained by area-level conditions. The findings are presented in **Table 6**. After adjusting for neighborhood characteristics, the lower obesity risk observed among foreign-born residents of East Asian enclaves remained significant (OR=0.55, *p* <.01), suggesting that this association is not explained by the measured community-level factors. In contrast, the elevated risks for diabetes and hypertension previously observed in South Asian enclaves were attenuated and no longer statistically significant after accounting for community-level factors. For diabetes, the OR decreased from 1.41 to 1.26 (an approximate 11% reduction), and for hypertension, the OR decreased from 1.50 to 1.27 (about a 15% reduction).

**Table 6.** Associations between Asian Enclave Residency and Cardiometabolic Outcomes Adjusting for Community-level Factors, Among Foreign-born.

|  | Obesity |  |  |  | Diabetes |  |  |  | Hypertension |  |  |  |
| --- | --- | --- | --- | --- | --- | --- | --- | --- | --- | --- | --- | --- |
|  | OR |  | 95% CI |  | OR |  | 95% CI |  | OR |  | 95% CI |  |
| **Enclave residency** |  |  |  |  |  |  |  |  |  |  |  |  |
| *Non-Enclaves (ref)* |  |  |  |  |  |  |  |  |  |  |  |  |
| *East Asian Enclaves* | 0.55 | ** | 0.36 | 0.83 | 0.99 |  | 0.68 | 1.44 | 1.01 |  | 0.73 | 1.41 |
| *South Asian Enclaves* | 1.33 |  | 0.94 | 1.88 | 1.26 |  | 0.93 | 1.71 | 1.27 |  | 0.95 | 1.71 |
| **Area characteristics** |  |  |  |  |  |  |  |  |  |  |  |  |
| Socioeconomic deprivation | 1.12 |  | 0.82 | 1.52 | 1.42 | * | 1.07 | 1.89 | 1.14 |  | 0.88 | 1.47 |
| Social cohesion | 1.18 |  | 0.85 | 1.65 | 1.21 |  | 0.94 | 1.56 | 1.02 |  | 0.82 | 1.28 |
| Built environment | 1.04 |  | 0.82 | 1.31 | 1.00 |  | 0.81 | 1.23 | 0.93 |  | 0.79 | 1.09 |
| Institutional support for immigrants | 1.09 |  | 0.87 | 1.37 | 0.98 |  | 0.83 | 1.14 | 0.89 | + | 0.78 | 1.02 |

\*\* $p < .01$ ; \* $p < .05$ ; + $p < .10$ .

### 4.9. Sensitivity Analysis

Since there are no universally established criteria for defining Asian enclaves, prior studies have applied concentration thresholds ranging from 5% to 20%^18,26,28,30^. Thus, to assess the robustness of our findings to different definitional choices, we tested several alternative enclave operationalizations: (1) a concentration-based definition using the double share threshold only (**S Figure 1** left panel), (2) a more stringent concentration-based definition using a triple share threshold only (**S Figure 1** right panel), (3) spatial clustering using local Moran’s I only (**S Figure 2**), and (4) a hybrid definition combining the triple share concentration with spatial clustering (**S Figure 3**). As presented in **S Table 2**, results were largely consistent across these definitions. For East Asian enclaves, most approaches continued to show lower odds of obesity, with no significant associations for diabetes or hypertension. South Asian enclave measures consistently indicated higher odds of diabetes and hypertension across operationalizations.

## 5. Discussion

This study offers new insight into how ethnic-specific neighborhood contexts shape CMB risks among AAs, revealing patterns that challenge the prior view of ethnic enclaves as uniformly health-protective. By disaggregating Asian populations and operationalizing enclaves at the ethnic subgroup level, we identify differentiated health risks that are obscured when using aggregated, pan-ethnic measures. Our findings also highlight how the relationship between enclave residence and health is shaped by nativity and can be partly explained by neighborhood-level conditions. In the sections that follow, we discuss our findings in relation to each of the three guiding research questions.

Our findings demonstrate a clear geographic distinction among ethnic-specific Asian enclaves in New York City, particularly between East Asian and South Asian communities. These groups occupy separate residential spaces, a pattern consistent with prior work documenting spatial separation across Asian subgroups^31,42^. This may help explain the largely mixed findings in prior studies that used pan-national Asian enclaves, which ranged from null effects on health^5,22^, to protective effects^7,20^, and detrimental effects^1,17^.

More importantly, our findings reveal that the health implications of enclave residence vary substantially across ethnic-specific Asian enclaves. East Asian enclaves appear protective against obesity, not only for East Asian individuals but also for other Asian subgroups. In contrast, South Asian enclaves are associated with increased risks for diabetes and hypertension among both South Asians and other Asian groups. These patterns are consistent with prior research highlighting the heterogeneity of enclave effects across ethnic-specific enclaves. For example, Janevic et al.^26^ reported elevated risk of gestational diabetes among women residing in South Asian enclaves in New York City, but found no such association in Chinese neighborhoods. These findings underscore that the health consequences of enclave residence are not uniform across AA populations, and the need to move beyond broad racial categories and toward ethnic-specific measures.

We also find that the health effects of ethnic enclave residence differ meaningfully by nativity. Protective associations between East Asian enclave residence and obesity, as well as the adverse associations between South Asian enclaves and CMB outcomes, were observed only among foreign-born Asians. Among US-born Asians, these associations were not significant. This pattern contrasts with prior research suggesting that enclaves may offer psychosocial benefits for immigrants while posing risks for US-born Asians^27^. Our findings may be better understood by differential sensitivity to neighborhood context, driven by differences in reliance on the immediate residential environments. Kalmijn^43^ found that immigrants tend to be more embedded in their neighborhoods than natives, in part due to greater demand for practical and social support arising from limited access to mainstream institutional resources and experiences of social exclusion in wider society. As a result, immigrants often rely on local resources to navigate unfamiliar institutions and fulfill daily needs, which can make them more susceptible to both the health-supportive and health-constraining features of enclave settings. In contrast, US-born Asians may have broader institutional familiarity and greater individual-level resources that reduce their dependence on the immediate neighborhood. It is also possible that differences in health domains contribute to these contrasting patterns. Prior study focusing on psychosocial outcomes may be capturing different mechanisms than those influencing CMB health risks. Future research should investigate how nativity interacts with neighborhood context across a wider range of health domains, and whether similar patterns of differential sensitivity emerge in other immigrant-origin populations.

Our findings reveal important similarities and differences in neighborhood conditions across ethnic-specific Asian enclaves. Both East Asian and South Asian enclaves experienced slightly higher levels of socioeconomic disadvantage than non-enclave areas, a pattern consistent with prior studies of urban Asian enclaves, which often contrast with more affluent suburban counterparts^28–30^. On the other hand, neighborhood social cohesion indicators were relatively similar between Asian enclaves and non-enclave neighborhoods. This diverges from longstanding theoretical perspectives, which posit that enclaves foster strong ethnic networks and institutions that offer emotional and instrumental support^35^, which can promote health through reduced exposure to discrimination and reinforcement of healthy behaviors stemming from the culture of origin^10,11^. However, several empirical studies have reported that AA enclave residents often perceive lower levels of social cohesion^16,18^ and social capital^17^. One possible explanation, as noted in prior research, is that the ethnic density of AAs is generally lower than that of other racial or ethnic groups, limiting the formation of cohesive networks large enough to generate collective benefits^16^. It is also important to note that our cohesion measures reflect general perceptions of neighborhood cohesion rather than cohesion specific among co-ethnic neighbors, thus potentially underestimating the types of social support most relevant within ethnic enclaves.

In addition, both Asian enclaves had worse built environment conditions than non-enclave areas, pointing to broader infrastructural underinvestment. These disparities are consistent with prior work. In Los Angeles County, Asian-majority neighborhoods receive significantly less park space per capita, just 1.2 acres per 1,000 residents, compared with 17.4 acres in white-majority areas^36^. These differences are not a result of a general lack of green space in the county but reflects how public resources are unevenly distributed across racial and ethnic lines^37^. The place stratification perspective^44^ offers a useful lens for understanding these patterns, highlighting how discriminatory practices in the housing market and unequal political representation contribute to sustained racial segregation and unequal neighborhood conditions, regardless of length of residency in the US and individual socioeconomic resources^11,45^. As a result, communities that lack the political or institutional capacity to mobilize resources are more likely to suffer from poor infrastructure and limited access to health-promoting amenities.

Enclaves differed in aggregated resident characteristics and the degree of institutional investment. East Asian enclave residents had higher rates of limited education and poverty, features aligned with early-stage immigrant communities^17,46^. Yet, East Asian enclaves received the highest levels of immigrant-targeted funding. In contrast, South Asian enclaves reflected a disconnect between individual-level advantage and neighborhood-level disinvestment. Despite residents’ relatively high educational attainment, these enclaves had the most under-resourced built environments and the lowest levels of immigrant-related funding. This mismatch aligns most closely with the place stratification framework, which emphasizes how racialized structures can prevent even socioeconomically mobile immigrant groups from translating individual gains into improved neighborhood conditions^11^.

Our measured neighborhood-level factors accounted for some, but not all, of the observed associations between ethnic enclave residence and CMB outcomes among foreign-born AAs. For East Asian enclaves, the protective association with obesity was largely unchanged even after adjusting for neighborhood socioeconomic disadvantage, social cohesion, built environment, and institutional support. This persistence suggests that the lower obesity risk is not attributable to the neighborhood characteristics captured in our data. It points to the presence of other unmeasured, enclave-specific pathways that are beneficial but not captured by our neighborhood-level measures. We elaborate on these potential mechanisms later in this section. For South Asian enclaves, the elevated risks for diabetes and hypertension were attenuated and became statistically non-significant once neighborhood factors were included. The OR for diabetes declined by 11%, and the OR for hypertension decreased by 15%. These shifts indicate that neighborhood conditions may contribute to the observed patterns; however, they explain only a small portion of the association and the loss of significance likely reflects, in part, reduced statistical precision after covariate adjustment. This pattern also suggests the relevance of additional mechanisms not captured in our neighborhood-level indicators.

One such pathway may involve cultural differences in health behaviors. Ethnic enclaves often facilitate access to culturally preferred foods and traditional diets, where greater availability of ethnic groceries and restaurants supports the retention of healthier eating patterns^7^. Studies suggest that East Asian diets, characterized by low intake of red meat and high intakes of fish and plant foods^47^ are similar to the Mediterranean diet^48^ and have been linked to cardiovascular health. As a result, East Asian enclaves likely promote healthier diets through restaurants and food options aligned with these traditional, low-fat, vegetable-rich meals, contributing to lower cardiometabolic risks. In contrast, South Asian diets are reported to have high intakes of carbohydrates, saturated fats, and omega-6 polyunsaturated fatty acids, with lower intake of monounsaturated fats and fiber, which are linked to obesity, insulin resistance, diabetes, and cardiovascular disease^49,50^. Compounding this, high levels of physical inactivity have been reported among South Asian immigrants in countries such as Australia^51^, Canada^52^, and the UK^53^, along with gendered barriers to exercise^50^ where women face norms related to religious modesty, avoidance of mixed-sex activities, and concerns about going out alone^54^. These cultural expectations often place family and household responsibilities above physical activity, limiting opportunities for women to engage in exercise^55^. These country-of-origin norms, when reinforced within enclaves, may reduce opportunities for health-promoting behaviors and contribute to increased CMB risk.

Another potentially important but unmeasured mechanism is the role of ethnic-specific cultural institutions. Prior studies suggest that the effects of enclaves may depend not only on the co-ethnic population density but also on the presence of robust co-ethnic businesses and services, including clinics, cultural centers, religious institutions, and schools, that can reduce experiences of discrimination and facilitate access to culturally congruent resources^27^. These institutions contribute to a strong “ethnic feel,” marked by visible cultural cues such as signage, language, and retail offerings^56^. For example, Chinese enclaves in California’s San Gabriel Valley have been described as institutionally complete and culturally vibrant^33,57^. Zhou and Kim^35^ similarly describe how Chinese and Korean neighborhoods often feature well-developed educational infrastructures, including ethnic language schools and after-school academies, that both reinforce cultural identity and support academic success. These types of institutions may promote upward social mobility, greater access to information, and network-building, mechanisms that help explain the protective associations we observed in East Asian enclaves. In contrast, fewer studies have documented similar levels of institutional development in South Asian enclaves. These differences in institutional capacity may further account for the elevated CMB risks observed in South Asian enclaves in our study. Although we were unable to measure these institutional differences directly, future research should examine whether variation in enclave infrastructure helps account for divergent health trajectories across Asian ethnic enclaves.

### 5.1. Limitations

Several limitations warrant consideration. First, the cross-sectional design limits our ability to draw causal inferences. Observed associations may reflect residential self-selection rather than enclave effects per se. Longitudinal data are needed to disentangle whether ethnic enclaves shape health trajectories over time or whether individuals with particular health profiles are more likely to reside in certain neighborhoods. Second, although our enclave definitions are more refined than pan-ethnic classifications, they are constrained by the geographic resolution at the PUMA-level, which may obscure important variation at finer spatial scales. Future research should leverage more granular geographic units, such as census tracts, to better capture neighborhood-level dynamics and heterogeneity within enclaves. Third, our neighborhood-level indicators may not fully capture residents’ experiences. Future studies should incorporate additional indicators that reflect features central to the health effects of ethnic enclaves, such as the density of ethnic businesses, cultural institutions, and community organizations, as well as food-environment measures (e.g., availability and accessibility of culturally specific groceries and restaurants). Finally, the generalizability of our findings is limited to New York City, a metropolitan area with distinctive patterns of Asian immigration and settlement. Replication in other contexts is needed to assess the broader applicability of these results.

### 5.2. Conclusion

Overall, our results suggest that residence in East Asian enclaves may be protective against obesity, while residence in South Asian enclaves may be associated with elevated risks of diabetes and hypertension among foreign-born AAs. These findings challenge the conventional view that Asian ethnic enclaves are universally protective for health. Our results underscore the need for health-specific, ethnicity-specific, and nativity-disaggregated approaches when examining the impact of Asian enclaves on health outcomes. By exploring distinct explanatory pathways, we highlight the importance of moving beyond traditional neighborhood factors, such as socioeconomic status and social cohesion, to incorporate dimensions like the built environment and the investment for immigrants. These multidimensional factors are critical for understanding the mechanisms through which enclaves shape health and for identifying pathways that may confer risk or resilience across diverse Asian subgroups.

## Data Availability

This study draws on both publicly available data sources and restricted data product from the New York City Community Health Survey, which is conducted by the Ney York City Department of Health and Mental Hygiene. Public-use files are freely downloadable from the website. Data on participants geographic location are available via a data use agreement with the NYC Department of Health. Community-level data used in this study are publicly available from the American Community Survey, NYC Plannings Community District Profiles, the NYC Department of Sanitation via the Environment & Health Data Portal, and Data2Go.NYC.

https://www.nyc.gov/site/doh/data/data-sets/community-health-survey.page

