## Supplemental materials for "Not One Enclave: Disaggregation and Cardiometabolic Health in Asian Ethnic Enclaves"

**List of Supplemental Materials**

**S Appendix 1**. Neighborhood Environment Measures

**S Table 1.** Factor Analyses of Neighborhood Indicators

**S Figure 1.** Geographic Locations of Ethnic-Specific Asian Enclaves in New York City

**S Figure 2.** Spatial Clustering Analysis

**S Figure 3.** Geographic Locations of Ethnic-Specific Asian Enclaves in New York City, Triple-share Criteria, Spatial Clustering Adjusted

**S Table 2.** Associations between Asian Enclaves and Health Outcomes among All Asians

**S Appendix 1.** Neighborhood Environment Measures

To assess neighborhood socioeconomic conditions and built environment features at PUMA-level, we initially examined 16 indicators: poverty rate, rent burden, percentage without a bachelor’s degree, unemployment rate, park access, number of hospitals, number of libraries, number of public schools, ratio of bodegas to supermarkets, number of public facilities, transportation or utility infrastructure, open space coverage, street cleanliness, vegetation coverage, proportion of very old housing, and litter basket coverage. We refined this set of indicators using exploratory factor analysis, excluding variables with low factor loadings (below 0.3). We retained eight variables for analysis: poverty rate, rent burden, no bachelor’s degree, unemployment, park access, number of hospitals, public facilities, and litter basket coverage. *The poverty rate* is based on the NYCgov poverty measure, a locally adjusted metric that accounts for the high cost of housing in New York City and incorporates non-cash benefits and expenses like medical care, commuting, and childcare. This measure provides a more realistic assessment of poverty in the city than the federal definition. *Rent burden* captures the percentage of households spending 35% or more of their income on rent. *Educational attainment* is measured by the percentage of residents without a bachelor’s degree, and *unemployment* reflects the proportion of the civilian labor force that is unemployed. *Park access* is defined as the percentage of residents living within walking distance, ¼ mile for small parks and ½ mile for larger parks and open space. The availability of *healthcare infrastructure* is represented by the number of hospitals, urgent care centers, diagnostic and treatment facilities, and school-based health centers. *Public facilities* are measured as the percentage of lot area dedicated to institutions or facilities with public use as their primary land function. Lastly, *litter basket coverage* is defined as the density of public trash cans per square mile, which supports cleaner public environments and may help reduce littering.

We conducted another factor analysis using these eight PUMA-level indicators to identify latent dimensions of neighborhood environment. As shown in **S Table 1A**, an oblimin rotation of a two-factor solution revealed that Factor 1 loaded strongly on poverty rate, rent burden, lack of a bachelor’s degree, and unemployment, reflecting neighborhood socioeconomic disadvantage. Factor 2 captured features of the built environment, with high loadings for park access, number of hospitals, public facilities, and litter basket density. Factor scores for each construct were predicted and used as continuous variables in subsequent regression models.

We also considered indicators of area-level social cohesion, drawing from responses to the NYCCHS and aggregating them to the PUMA level. Because data on these items were not collected consistently across all years of our analytic sample (2015–2020), we used responses from the years in which each item was available. Available items reflect perceptions of neighborhood connectedness, including whether residents feel they live in a close-knit neighborhood (2015–2016), believe people are willing to help their neighbors (2015–2018), perceive that people in the neighborhood get along (2015–2016), feel that people share the same values, and trust others in the neighborhood (2015–2016). Each indicator represents the percentage of adults aged 18 and older who reported that they “strongly agree” or “somewhat agree” with the corresponding statement. To create a composite measure of neighborhood social cohesion, we conducted a factor analysis on five items. As shown in **S Table 1B**, a one-factor solution was clearly supported (eigenvalue=3.40), accounting for 93.8% of the common variance. All five items loaded positively on this factor (all loadings > 0.30). Factor scores were predicted and used as a continuous measure of neighborhood cohesion.

Finally, we included a single measure of public funding allocated for immigrant family at each PUMA. The measure reflects budget allocations supporting immigrant communities through programs including civic classes and English for Speakers of Other Languages, immigration application assistance.

**S Table 1.** Factor Analyses of Neighborhood Indicators

A. Neighborhood Socioeconomic Status and Built Environment

| **Variable** | **Factor 1** | **Factor 2** |
| --- | --- | --- |
|  | Socioeconomic Status | Built Environment |
| Poverty rate | .88 | .21 |
| Rent burden rate | .83 | -.23 |
| % without bachelor’s degree | .92 | -.28 |
| Unemployment rate | .70 | .36 |
| Park access | -.01 | .76 |
| Number of hospitals | .15 | .71 |
| % of area as public facilities | -.02 | .59 |
| Litter basket density | -.52 | .72 |
| Eigenvalue | 3.12 | 2.25 |
| % of common variance explained | 58.7 | 42.4 |

B. Neighborhood Social Cohesion

| **Variable** | **Factor 1** |
| --- | --- |
| People in this neighborhood are close-knit | .30 |
| People are willing to help each other | .91 |
| People get along with each other | .90 |
| People share the same values | .91 |
| People in this neighborhood can be trusted | .92 |
| Eigenvalue | 3.40 |
| % of common variance explained | 93.84 |

**S Figure 1.** Geographic Locations of Ethnic-Specific Asian Enclaves in New York City

| **A. East Asian Enclave** | |
| --- | --- |
| A1. Double share (16.4%) (n=9) | A2. Triple share 24.6% (n=5) |
| 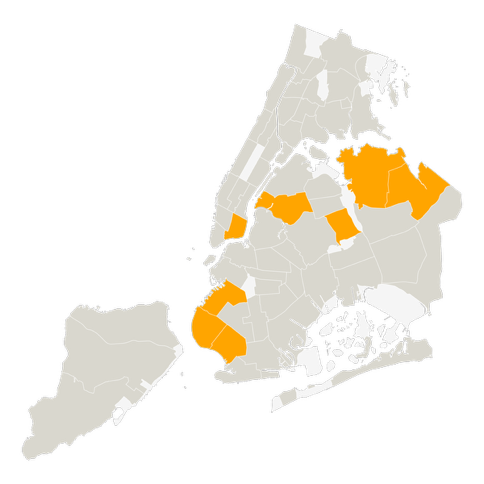 | 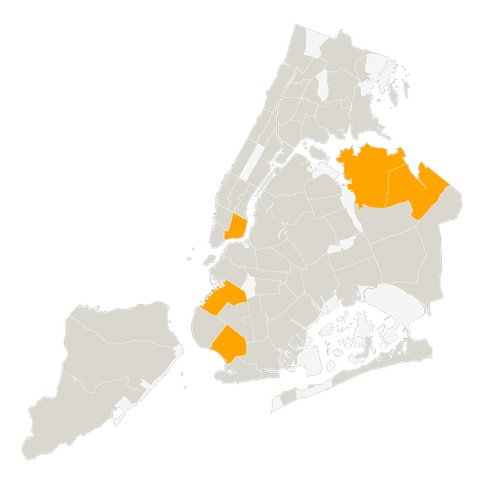 |
| **B. South Asian Enclave** | |
| B1. Double share (8.6%) (n=7) | B2. Triple share (12.9%) (n=3) |
| 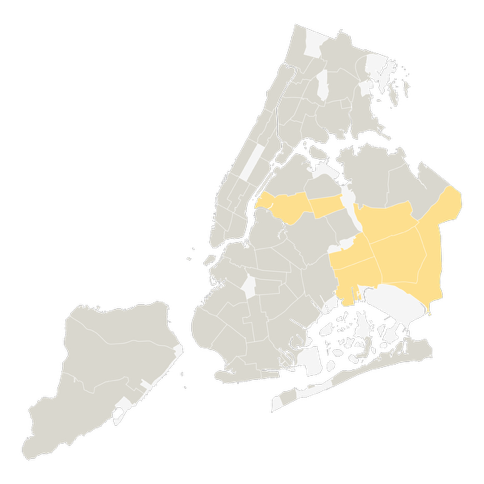 | 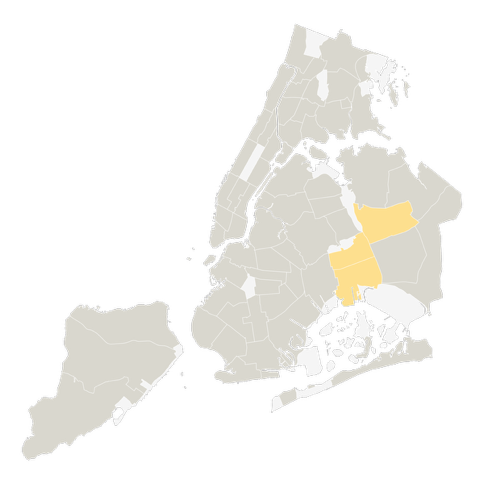 |

**S Figure 2. Spatial Clustering Analysis**

| **A1. East Asian tracts** | **A2. East Asian aggregated at PUMA** |
| --- | --- |
| 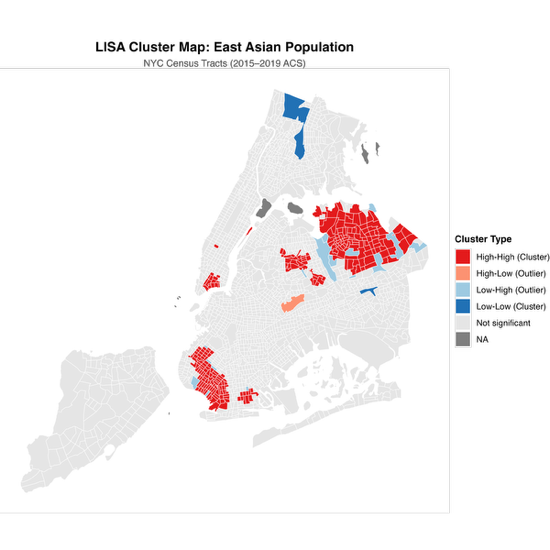 | 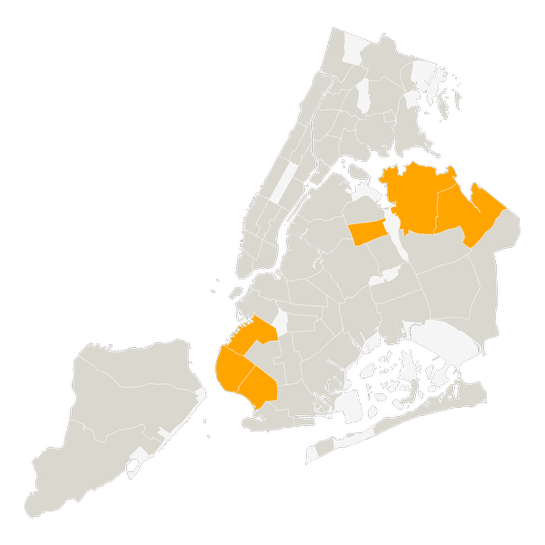 |
| **B1. South Asian tracts** | **B2. South Asian aggregated at PUMA** |
| 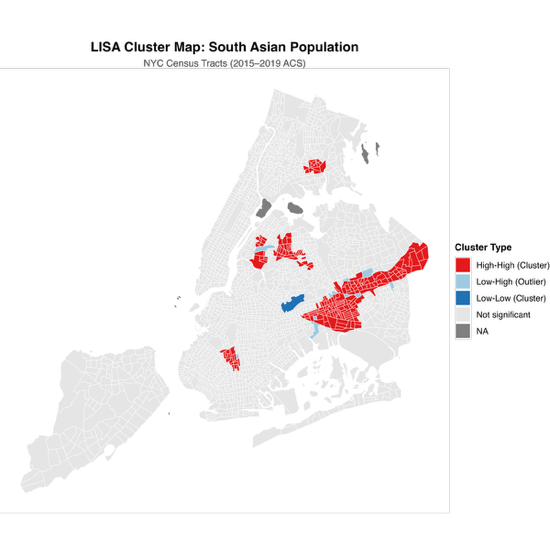 | 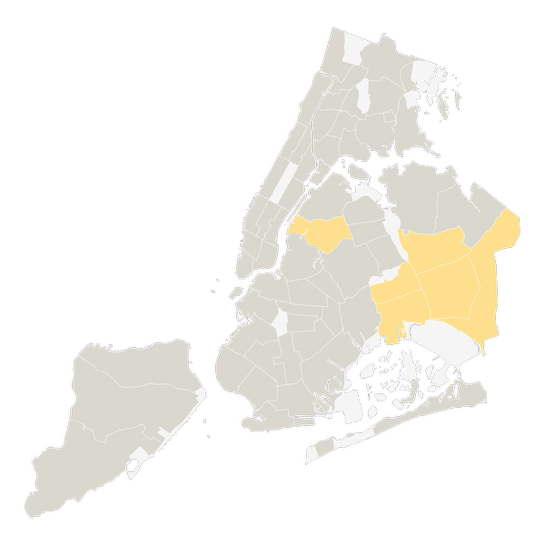 |

**S Figure 3.** Geographic Locations of Ethnic-Specific Asian Enclaves in New York City, Triple-share Criteria, Spatial Clustering Adjusted

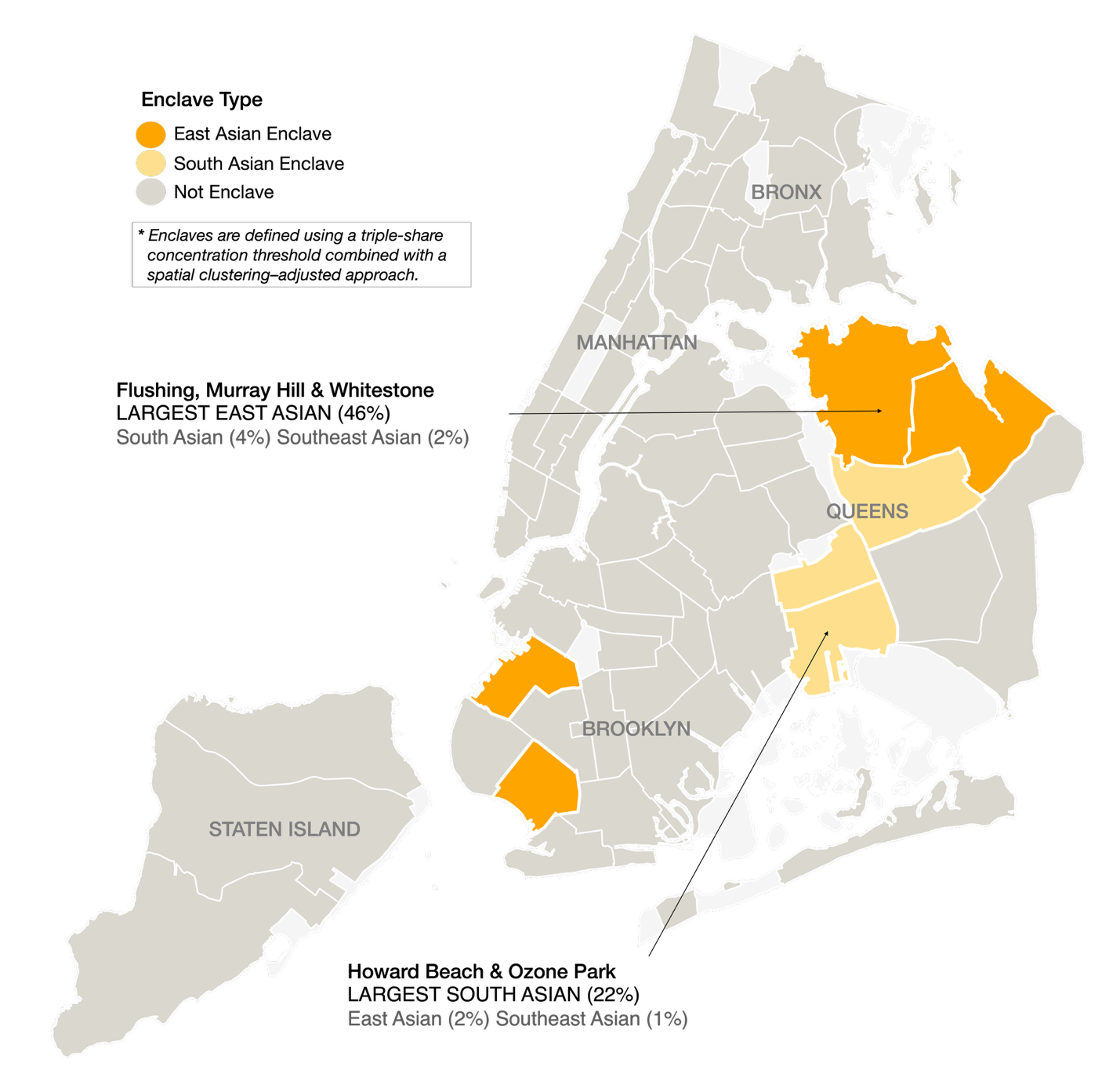

*Note.* Light grey areas are joint interest areas such as parks and airports.

**S Table 2.** Associations between Asian Enclaves and Health Outcomes among All Asians

|  | **Obesity** | | | | **Diabetes** | | | | **Hypertension** | | | |
| --- | --- | --- | --- | --- | --- | --- | --- | --- | --- | --- | --- | --- |
|  | **OR** |  | **95% CI** | | **OR** |  | **95% CI** | | **OR** |  | **95% CI** | |
| ***East Asian Enclave*** |  |  |  |  |  |  |  |  |  |  |  |  |
| Double-share, spatial clustering both | 0.63 | * | 0.41 | 0.96 | 1.03 |  | 0.78 | 1.35 | 1.06 |  | 0.79 | 1.42 |
| Double-share only | 0.63 | * | 0.44 | 0.90 | 0.94 |  | 0.72 | 1.22 | 0.99 |  | 0.73 | 1.34 |
| Triple-share only | 0.59 | ** | 0.40 | 0.87 | 0.91 |  | 0.72 | 1.17 | 0.90 |  | 0.67 | 1.21 |
| Spatial clustering only | 0.73 |  | 0.47 | 1.12 | 0.90 |  | 0.67 | 1.21 | 1.00 |  | 0.75 | 1.33 |
| Triple-share, spatial clustering both | 0.60 | * | 0.38 | 0.94 | 0.94 |  | 0.72 | 1.22 | 1.01 |  | 0.75 | 1.36 |
| ***South Asian Enclave*** |  |  |  |  |  |  |  |  |  |  |  |  |
| Double-share, spatial clustering both | 1.26 |  | 0.92 | 1.72 | 1.42 | * | 1.06 | 1.89 | 1.46 | * | 1.09 | 1.95 |
| Double-share only | 1.20 |  | 0.89 | 1.62 | 1.16 |  | 0.82 | 1.65 | 1.26 |  | 0.91 | 1.73 |
| Triple-share only | 1.15 |  | 0.73 | 1.81 | 1.43 | ** | 1.15 | 1.79 | 1.33 | ** | 1.09 | 1.61 |
| Spatial clustering only | 1.26 |  | 0.92 | 1.74 | 1.37 | * | 1.03 | 1.81 | 1.44 | * | 1.07 | 1.93 |
| Triple-share, spatial clustering both | 1.16 |  | 0.74 | 1.82 | 1.44 | *** | 1.17 | 1.79 | 1.36 | ** | 1.12 | 1.66 |
| **Observations** | 6354 |  |  |  | 6436 |  |  |  | 6434 |  |  |  |

*Note.* All models were adjusted for age group, sex, Asian ethnic subgroup membership, nativity, marital/partner status, educational attainment, household income, and health insurance status.

*** *p* < .001; ** *p* < .01; * *p* < .05.
